# Persistently reduced humoral and cellular immune response following third SARS-CoV-2 mRNA vaccination in anti-CD20-treated multiple sclerosis patients

**DOI:** 10.1101/2022.01.27.22269944

**Authors:** Hamza Mahmood Bajwa, Frederik Novak, Anna Christine Nilsson, Christian Nielsen, Dorte K. Holm, Kamilla Østergaard, Agnes Hauschultz Witt, Keld-Erik Byg, Isik S. Johansen, Kristen Mittl, William Rowles, Scott S. Zamvil, Riley Bove, Joseph J. Sabatino, Tobias Sejbaek

**Author notes:** These authors contributed equally (Hamza Mahmood Bajwa, Frederik Novak). Pre-graduate fellow H. M. Bajwa, Dr. Novak, & Dr. Sejbaek conducted the statistical analysis and graphical presentations in collaboration. Corresponding author: Dr. Tobias Sejbaek, Department of Neurology, Hospital of South West Jutland, Esbjerg, Denmark, Finsensgade 35, 6700 Esbjerg, Denmark.

## Abstract

**Objective:** To examine humoral and cellular response in multiple sclerosis patients on anti-CD20 therapy after third BNT162b2 mRNA SARS-CoV-2 vaccination.

**Methods:** A prospective longitudinal study design from first throughout third vaccination in Danish and American MS centers. All participants were treated with ocrelizumab. Antibody (Ab) levels were assessed before and after third vaccination using SARS-CoV-2 IgG II Quant assay (Abbott Laboratories). B- and T-lymphocytes enumeration was done with BD Multitest™6-color TBNK reagent. Spike-specific T-cell responses were measured through PBMC stimulation with spike peptide pools (JPT Peptide Technologies).

**Results:** We found that 14.0%, 37.7%, and 33.3% were seropositive after first, second and third vaccination. The median Ab-levels were 74.2 BAU/mL (range: 8.5-2427), 43.7 BAU/ml (range: 7.8-366.1) and 31.3 BAU/mL (range: 7.9-507.0) after first, second and third vaccination, respectively. No difference was found in levels after second and third vaccination (p=0.1475). Seropositivity dropped to 25.0% of participants before the third vaccination, a relative reduction of 33.3% (p=0.0020). No difference was found between frequencies of spike reactive CD4^+^ and CD8^+^ T-cells after second (0.65 ± 0.08% and 0.95 ± 0.20%, respectively) and third vaccination (0.99 ± 0.22% and 1.3 ± 0.34%), respectively.

**Conclusion:** In this longitudinal cohort we found no significant increased humoral or cellular response with administration of a third SARS-CoV-2 mRNA vaccination. These findings suggest the need for clinical strategies to include allowance of B cell reconstitution before repeat vaccination and/or provision of pre-exposure prophylactic monoclonal antibodies.

**Key Points:** *What is already known on this topic:* Studies have described decreased humoral response and sustained T-cell reactivity after standard two-dose SARS-CoV-2 mRNA vaccination during anti-CD20 therapy in multiple sclerosis participants.

*What this study adds:* Persistently decreased humoral, but stable cellular reactivity following a third SARS-CoV-2 mRNA vaccination.

*How this study might affect research, practice or policy:* The findings suggest the need for clinical strategies to include allowance of B cell reconstitution before repeat vaccination and/or provision of pre-exposure prophylactic monoclonal antibodies.

## Introduction

The ongoing Covid-19 pandemic raises concerns about its effects on the most vulnerable patients. Anti-CD20 medications such as ocrelizumab, rituximab, and ofatumumab are widely used to treat multiple sclerosis (MS), blood cancers, and autoimmune diseases. Anti-CD20 targets B-lymphocytes, thus leading to cell lysis, and thereby reduction of disease activity in both relapsing and progressive MS (1). Furthermore, compared to other disease modifying therapies, anti-CD20 treatment is associated with more severe complications to SARS-CoV-2 infection (2) e.g., higher rates of hospitalizations and severe disease course (3). A COVID-19-specific strategy for patients at specific risk in Denmark has therefore been to offer early re-vaccination to anti-CD20 treated patients.

It has previously been shown by our and other investigators that patients on B-cell depleting treatments have significantly reduced humoral immunity after COVID-19 vaccines compared to healthy controls (4, 5). Several studies confirm that vaccination, in general, generates a decreased humoral response in anti-CD20 treated patients (6–9). Data indicate that higher levels of B-cells at the time of vaccination and longer intervals between anti-CD20 treatments improve the response to vaccination (10). However, extending dosage interval is currently considered an off-label treatment, and the effect on relapse risk, while appearing to date to not be substantial, is still unknown (9, 11–13). One study has advocated that vaccination should occur one month before the next treatment infusion (14). Still, the timing of treatment with anti-CD20 infusion and subject vaccination is still being debated (4, 15).

The initial two vaccinations can lead to successful seroconversion in a subset of anti-CD20 patients (4). Accordingly, the Danish National Board of Health, European Medicines Agency (EMA), and the U.S. Food and Drug Administration (FDA) have all recommended or authorized an additional third vaccine for these patients.

The primary goal of this study is to determine whether additional mRNA SARS-CoV-2 vaccination can increase levels of specific SARS-CoV-2 spike receptor binding domain (RBD) antibodies (Abs) generated in MS patients treated with anti-CD20 therapy (ocrelizumab). We also assessed whether a third vaccine dose can increase T cell responses and the proportion of seropositive individuals among these participants. To address this question, we examined frequencies of spike-reactive T cells and levels of SARS-CoV-2 Abs before and after a third SARS-CoV-2 vaccination in a large cohort of anti-CD20-treated MS-patients from two international MS centers.

## Method

### Study population and design

In this observational study, we included adult participants (18 years or older) with confirmed MS (2017 McDonald Criteria) on ocrelizumab (anti-CD20) therapy. All participants had received two doses of mRNA SARS-CoV-2 vaccination and were enrolled prior to a third booster vaccine of the mRNA SARS-CoV-2 vaccine. Results from first and second vaccination were already published in a recent paper (4).

No other immunosuppressive treatment beyond infusion-related methylprednisolone was given to the participants during this study. Patients were included from two Danish MS clinics (Esbjerg, Viborg) and the University of California, Centre for MS and Neuroinflammation, in San Francisco (USA). All participants followed standard clinical practice by their treating neurologist and interval of time between second and third vaccination was not standardised(16, 17).

### Sample collection

Blood samples were collected at two time points: 0–7 days before the third booster (V4) vaccination and two to four weeks after the third booster vaccination (V5).

Abs were compared with levels 0-7 days before the first vaccination (V1), 0-7 days before the second booster vaccination (V2), and 2-4 weeks after the second booster vaccination (V3) (4). Danish participants provided samples at all timepoints (V1-V5); North American participants at V1, V3, V4, and V5 **(Figure 1)**.

**Figure 1.**
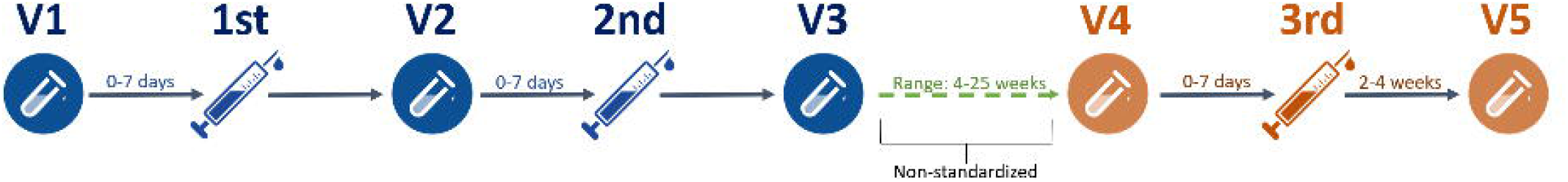

### Data collection

Blood samples were collected following international guidelines for biobanking. Venous blood was procured from a cubital vein into evacuated K2-EDTA or heparinized tubes. Plasma was aliquoted in 500 μL Sarstedt polypropylene tubes and stored at −80 °C until batch analysis (18).

#### Human Peripheral Blood Mononuclear Cell Isolation

Peripheral blood mononuclear cells (PBMCs) were isolated from whole blood using density gradient centrifugation (<u>Lymphoprep™</u>) and cryopreserved using DMSO-containing freezing medium and stored at – 196 °C until flow cytometric analyses

### Antibody assay and flow cytometry

We measured IgG antibodies against the SARS-CoV-2 spike RBD in plasma samples as described by Novak et al. (4) using the SARS-CoV-2 IgG II Quant assay (Abbott Laboratories), which is a quantitative chemiluminescent microparticle immunoassay (19). The assay was performed using the Abbott Alinity I platform in accordance with the manufacturer’s instructions. This assay has shown excellent correlation with the first WHO (World Health Organization) International Standard for anti-SARS-CoV-2 immunoglobulin (NIBSC code 20/136) (20), enabling the issuing of immunogenicity results in standardized units; binding antibody units (BAU)/mL for a binding assay format as the SARS-CoV-2 IgG II Quant assay. The mathematical relationship of the Abbott AU/mL unit to the WHO BAU/mL unit follows the equation BAU/mL = 0.142xAU/mL, corresponding to a cut-off at 7.1 BAU/mL. This assay has documented the ability to detect spike RBD IgG vaccine response in longitudinal samples from individuals with and without prior SARS-CoV-2 infection (19, 21). Ab-levels above 506 BAU/ml were defined as sufficient levels (>80% protection). Values between <506 BAU/ml and >17 BAU/ml were considered intermediate (>50% protection) and below <17 BAU/ml as low (22).

Enumeration of B- and T-lymphocytes was performed using fresh EDTA blood stained from the Denmark cohort with the BD Multitest™ 6-color TBNK reagent in BD TruCount tubes. Samples were analyzed on a BD FACSCanto™ II flow cytometer with BD FACSDiva software. Flow cytometry was performed in participants included from Hospital Southwest Jutland, University Hospital of Southern Denmark within 24 hours from collection.

### T cell assay

We measured spike-specific CD4+ and CD8+ T cell responses by activation-induced marker (AIM) expression as previously described(5). Briefly, PBMCs were thawed, washed, and rested for 2 hours at 37°C prior to start of assay. PBMCs were stimulated for 24 hours with 1 µg/ml spike peptide pools (JPT Peptide Technologies) or vehicle control (0.2% DMSO). Folllowing stimulation, cells were washed and stained with the following antibody panel: CD4 Alexa 488 (OKT4), CD8 Alexa 700 (SK1), OX-40 PE-Dazzle 594, (ACT35), CD69 PE (FN-50), CD137 BV421 (4B4-1), CD14 PerCP-Cy5.5 (HCD14), CD16 PerCP-Cy5.5 (B73.1), CD19 PerCP-Cy5.5 (HIB19) (all BioLegend) and live/dead dye eFluor506 (Invitrogen). Cells were washed with FACS wash buffer, fixed with 2% paraformaldehyde (BD), and stored in FACS wash buffer in the dark at 4°C until ready for flow cytometry analysis. The same gating strategy was employed as before(5). The frequencies of spike-specific T cells were calculated by subtracting the no stimulation background from spike peptide pool-stimulated samples.

### Standard protocol approvals, registrations, patient consents, and monitoring

Both written and oral consent was taken from all participants prior to inclusion. The Danish study followed national laws adhering to good clinical practice and was approved by the Danish National Committee on Health Research Ethics (Protocol no. S-20200068C) and Danish Data Protection Agency (journal no. 20/19,878). The study conducted by UCSF was made with the approval of the institutional review board (University of California, San Francisco, Committee on Human Research, protocol # 21–33240).

## Data availability

Anonymized data will be shared on request from any qualified investigator under approval from the Danish Data Protection Agency.

## Statistical analysis

All data were analyzed for normal distribution, and continuous data were presented as the median with minimum and maximum values. Mann-Whitney test was used for comparison between groups in case of missing values for paired analysis, and a Wilcoxon matched pairs signed ranks test was used for pairwise comparisons. Level of significance was defined as p<0.05.

### Results

We analysed IgG antibodies against the SARS-CoV-2 spike RBD in participants before and after the first, second, and third BNT162b2 vaccination. Samples before and after the third booster vaccination were collected between September 13 and November 25, 2021. All participants were vaccinated with BNT162b2. The median patient age was 47 years (range 24 - 67 years). Participant baseline demographic and clinical characteristics are shown in **Table 1**.

**Table 1.**
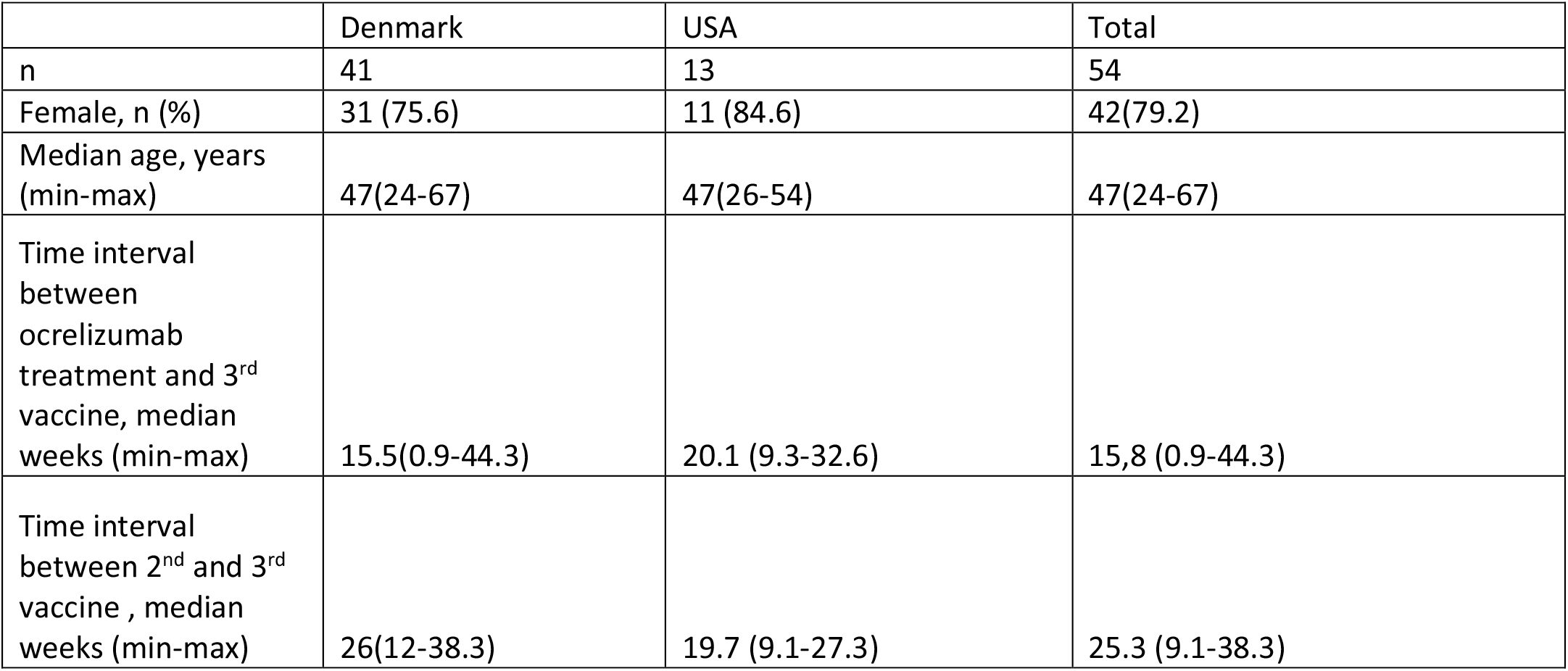
Baseline clinical and demographic characteristics.

|  | Denmark | USA | Total |
| --- | --- | --- | --- |
| n | 41 | 13 | 54 |
| Female, n (%) | 31 (75.6) | 11 (84.6) | 42(79.2) |
| Median age, years (min-max) | 47(24-67) | 47(26-54) | 47(24-67) |
| Time interval between ocrelizumab treatment and 3<sup>rd</sup> vaccine, median weeks (min-max) | 15.5(0.9-44.3) | 20.1 (9.3-32.6) | 15,8 (0.9-44.3) |
| Time interval between 2<sup>nd</sup> and 3<sup>rd</sup> vaccine , median weeks (min-max) | 26(12-38.3) | 19.7 (9.1-27.3) | 25.3 (9.1-38.3) |

#### Antibody levels before third vaccination

We found detectable Abs in 14.0% of participants at V2 (n=43), which increased to 37.7% at V3 (n=53) **(Fig. 2 C)**(4). At V4 (n=50), the frequency of seropositive patients decreased to 24.0% with a median level of 43.7 BAU/ml (range: 7.8-366.1 BAU/ml). 33.3% of patients with detectable Abs at V3 no longer had detectable Abs at V4, with their median level of Abs at V3 being 13.8 BAU/mL (range: 8.5-70.1). The patients with detectable Abs at both visits had a median Abs level of 276.5 BAU/mL (range: 22.8-2405 BAU/mL).The median time from V3 to V4 was 21.8 weeks (range: 4.1-25.1 weeks) (**Fig.1**) and the median time from V3 to V4 in participants who converted to negative was 20.3 weeks (range: 7.7-24 weeks). Overall, the Ab levels were significantly lower at V4 compared to V3 (p = 0.0020) **(Fig. 2 C)**.

**Figure 2.**
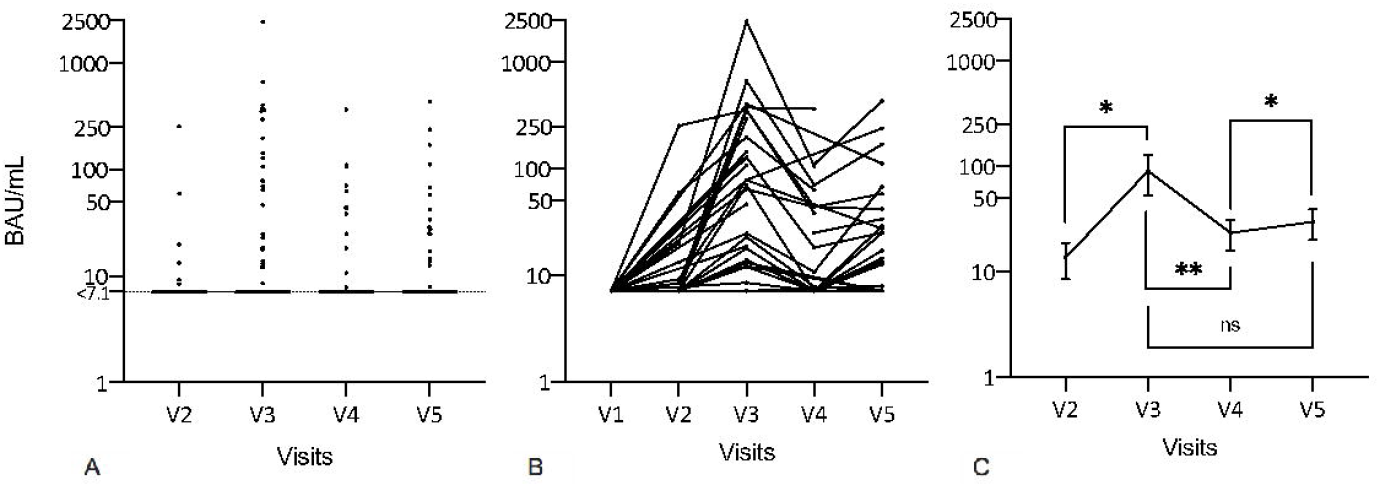

#### Antibody levels after the third vaccination

After third SARS-CoV-2 vaccination (V5), 18 out of 54 (33.3%) participants demonstrated positive Abs with a median of 31.3 BAU/mL (range: 7.9-507.0 BAU/mL). Paired results between V3 and V5 were available in 43 participants, of whom 11 out of 43 (25.6%) had detectable Abs at both V3 and V5 with a median of 41.8 BAU/mL (range: 12.6-238.5 BAU/mL). However, the proportion of seropositive patients between V3 and V5 was not significantly different (p= 0.1475) (**Fig.2 C**). Furthermore, 53 participants had paired results between V3/V4 and V5. In participants with previous undetectable AB-levels at V3/V4, 7 out of 53 (13.2%) participants had detectable Ab at V5 with a median of 14.4 BAU/mL (range: 7.9-18.8 BAU/mL). Mean Ab levels were significantly higher at V5 compared to V4 (p = 0.0313) (**Fig.2 C**).

#### Antibodies levels of protection after third vaccine (at V5)

When we categorized Ab levels according to protection against infection, 7.4% (n=4 out of 54) participants had low Ab levels between 7.1 and 17.0 BAU/ml, that would thereby be considered as providing less than 50% protection. In the intermediate group, 24.1% (n=13) had levels between 17-506 BAU/ml and are considered to have above 50% protection. One (1.8 %) patient had more than 506 BAU/ml and is considered to have protection >80%.

### Clinical and demographic predictors of Ab detectability

#### B-cells and T-cells

We compared levels of B-cells and T-cells between participants with or without detectable Abs at V4. No correlation were found in the levels of B-cells, CD4^+^ and CD8^+^ T lymphocytes between the seronegative and seropositive individuals.

#### Age, time between ocrelizumab infusion and third vaccination, and time from second to third vaccination

There was no difference in median age between seropositive (45 years, range: 26-58) (47 years, range: 24-67) participants (p= 0.2254 In addition, there was no difference between seronegative and seropositive participants in median time interval since the last infusion with ocrelizumab and the timing of third vaccination (15.7 weeks, range: 0.9-26.9 vs. 15.9 weeks, range: 3-44.3, n=52). Linear regression analysis revealed no correlation (r^2^= 0.04008, p= 0.44) between Abs level and the time between ocrelizumab infusion and third booster vaccine **(Fig. 3)**.

**Figure 3.**
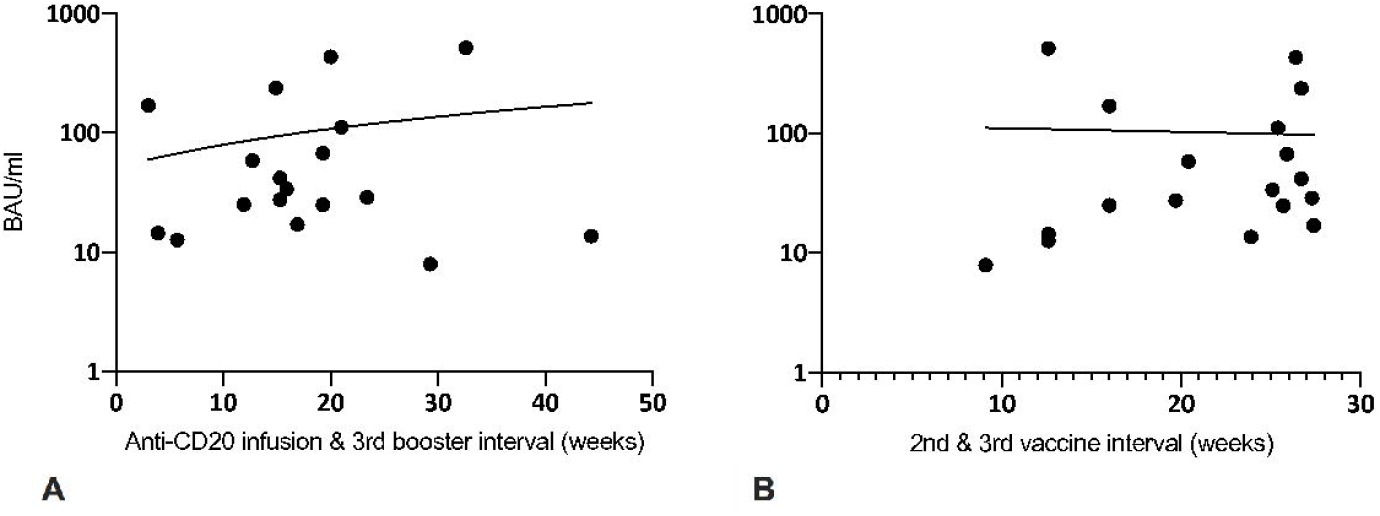

Finally, we compared levels of Abs and the time interval between the second and the third vaccination. There was no significant difference in median time intervals between seropositive (24.5 weeks, range: 9.1-27.4) and seronegative (26.0 weeks, range: 12-38.3), participants. In addition, we found no correlation between Abs level and time from second to third vaccination (r^2^=0.00097, p=0.90) **(Fig. 3)**.

#### Spike-specific CD4+ and CD8+ T cells

We also measured the frequencies of spike-reactive T cells at V1 (n=28), V3 (n=31) and V5 (n=23). Consistent with prior reports, the mean frequencies spike-specific CD4^+^ and CD8^+^ T cells at V3 (0.65 × 10^9^ cell/l ± 0.08% and 0.95 × 10^9^ cell/l ± 0.20%, respectively) were significantly increased compared to V1 (0.02 × 10^9^ cell/l ± 0.01% and 0.06 × 10^9^ cell/l ± 0.02%, respectively). While there was a marginal increase in spike-specific CD4+ and CD8+ T cell frequencies at V5 (0.99 × 10^9^ cell/l ± 0.22% and 1.3 × 10^9^ cell/l ± 0.34%, respectively), this was not significantly increased compared to V3 (**Fig. 4**).

**Figure 4.**
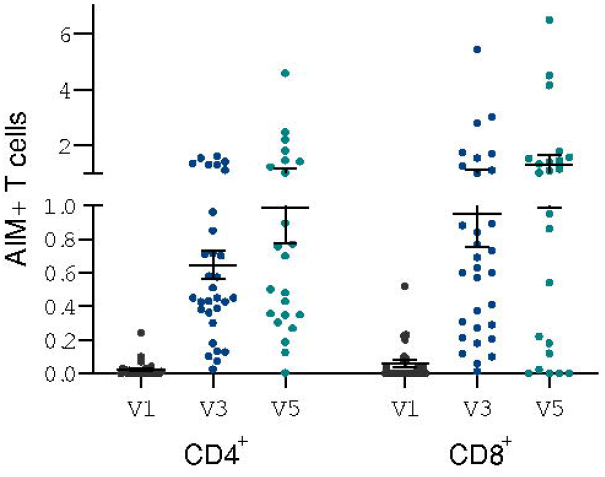

## Discussion

In this cohort of patients treated with ocrelizumab before mRNA SARS CoV-2 vaccination, we found minimal change in seroreactivity following a third vaccination, both in terms of seroconversion and antibody titers compared to healthy controls (23). In addition, the timing of anti-CD20 infusion and a third vaccination did not impact the antibody response.

We previously demonstrated that the majority of ocrelizumab treated patients converted from negative to positive Abs after the second vaccination. Thus, we expected a higher proportion of seropositive patients after the third vaccination (4) **(Figure 1)**. Here we compared proportions of SARS-CoV-2 Abs-positive participants after the first, second, and third BNT162b2 mRNA SARS-CoV-2 vaccine. We found that 14.0%, 37.7%, and 33.3% were seropositive, respectively. Seropositivity dropped to 25.0% of participants before the third vaccination, which is likely related to low antibody titers at V3 and to the median time of 25 weeks between the second and third vaccination (**Table 1**). To prevent conversion from positive to negative, a shortened interval between repeat vaccinations may be indicated in B-cell depleted patients.

A total of 26.7% participants in the seropositive group at V5 had no detectable levels of Abs at the previous visits (V1-V4), suggesting a certain degree of humoral reactivity can be achieved even in previously seronegative patients. Conversely, 20% of participants showed fluctuating seropositivity, converting from seropositivity at V3, to seronegativity at V4 and again to seropositivity at V5. These participants had low levels of Abs at V3, likely accounting for the fluctuating seropositivity. The same tendency is not observed in healthy controls (24–26). This suggests that even in those patients who achieve low titer seroconversion, the protective effects may be transient at best. The remaining proportion of the group was seropositive throughout V3-V5.

Our study did not find a correlation between anti-spike RBD Ab-levels and the time interval between vaccinations. More studies with different strategies, such as additional vaccination or permitting B cell reconstitution, are needed to optimize humoral responses to SARS-CoV-2 vaccination.

Several recent studies on third SARS-CoV-2 vaccine responses in anti-CD20 treated patients revealed seroconversion in a subset of individuals who had remained seronegative after two vaccinations (27–29). In contrast, our study of a longitudinal cohort from two international sites demonstrates that both the proportion of patients with positive Abs and the Ab-levels are overall unchanged compared to after the initial two dose vaccination.

Other studies have suggested that an extended dosage interval of ocrelizumab could improve the humoral response (10, 30). Extending interval is however controversial in regards of efficacy and not standard practice in Denmark and USA since the risk of disease activity with extended treatment interval is unknown. We found no correlation between levels of B-and T-cells and Ab-levels. B-cells are related to humoral response, and other studies have proven relationship between levels of B-cells and SARS-CoV-2 Abs **(Figure 2)**(10). Currently, the Danish Health Authority recommends the third vaccination 4.5 months after the second vaccination and US Centers for Disease Control and Prevention (CDC) recommends additional vaccine shots to be given after 5 months from third vaccination. Such recommendation will result in re-vaccination more than twice a year and extended treatment intervals in this scenario is logistically not possible and could be harmful to the patient.

Our study also assessed T cell responses following third SARS-CoV-2 vaccination in anti-CD20-treated MS patients. While spike-specific CD4+ and CD8+ T responses remained high after third vaccination, they were not significantly increased compared to the responses after the second vaccination. These findings therefore suggest that additional vaccinations do not significantly augment the cellular response in anti-CD20-treated patients.

There were limitations to our study. The short inclusion timeframe resulted in a few missing samples at V4 and V5. Furthermore, the cut-off levels presented in this study reflect reactivity against alpha variants of SARS-CoV-2, but not more recent variants of concern, including the delta or omicron variants.

In summary, in contrast to prior studies, in this longitudinal cohort we found no significant increased protective benefit from a humoral or cellular perspective with administration of a third SARS-CoV-2 mRNA vaccination. These findings have important clinical implications and suggest the need for clinical strategies to include allowance of B cell reconstitution before repeat vaccination and/or provision of pre-exposure prophylactic monoclonal antibodies.

## Acknowledgments

We acknowledge the great help received from patients participating in this trial and our team of technicians and study coordinators represented by Gunhild Brixen^1^ Nielsen, Sarah Andersen^1,^ and Pia Hannesbo^6^. We acknowledge grants given by the Lundbeck Neurological Scholarship delegated by the Danish Neurological Society and by Roche.

## Author contributions

H Bajwa, F Novak, T Sejbaek, J Sabatino, I S Johansen, KE Byg, and R Bove all contributed to study design

H Bajwa, F Novak, T Sejbaek, J Sabatino, W Rowles, AH Witt, K Østergaard all contributed to patient recruitment and patient data

F Novak, T Sejbaek, J Sabatino, A C Nilsson, C Nielsen, K Mittl, and W Rowles all contributed to sample processing

H Bajwa, F Novak, T Sejbaek, J Sabatino, A C Nilsson, C Nielsen, I S Johansen, KE Byg, AH Witt, K Østergaard, S Zamvil, and R Bove all contributed to data analysis/interpretation

