## Supplementary material for "Persistently reduced humoral and cellular immune response following third SARS-CoV-2 mRNA vaccination in anti-CD20-treated multiple sclerosis patients": Figure 1 legend

Sample collection at visits (V_x_) and vaccinations. V1= Visit 1 (0-7 days before the 1st vaccine), V2= Visit 2 (0-7 days before the 2nd vaccine), V3 = Visit 3 (2-4 weeks after the 2nd vaccine dose), V4 = Visit 4 (0-7 days before the 3rd booster) and V5 = Visit 5 (2-4 weeks after the 3rd booster). 1st, 2nd, and 3rd depict the order of vaccination. The time interval between V3 and V4 is based on standard clinical practice when patients were offered the 3^rd^ booster vaccine.
