## Supplementary material for "Persistently reduced humoral and cellular immune response following third SARS-CoV-2 mRNA vaccination in anti-CD20-treated multiple sclerosis patients": Figure 2 legend

Depicts levels of antibodies (Abs) given in BAU/mL- Y-axis with logarithmically scaled. BAU/ml = Binding antibody unit per ml. V1= Visit 1 (0-7 days before the 1st vaccine), V2= Visit 2 (0-7 days before the 2nd vaccine), V3 = Visit 3 (2-4 weeks after the 2nd vaccine dose), V4 = Visit 4 (0-7 days before the 3rd booster vaccine) and V5 = Visit 5 (2-4 weeks after the 3rd booster). Cut-offs are depicted with dotted lines. We defined seronegative (undetectable Abs) as < 7.1 BAU/mL, low levels of Abs as 7.1-17 BAU/mL, intermediate levels of Abs as 17-506 BAU/mL and high levels of Abs as > 506 BAU/mL.

Graph A: Ab-Levels after 1^st^ vaccination and before 2^nd^ vaccine at V2, after 2^nd^ vaccine at V3, before 3^rd^ booster at V4 and after the 3^rd^ booster at V5. Each dot corresponds to the measured Ab-level.

Graph B: Outlines the Ab-development from V1-V5. All patients at V1 were seronegative.

Graph C: Depicts the mean of levels of Abs at each visit with standard error of mean. Lines demonstrates levels of significance (p-value) between visits in paired samples V2 vs V3 (p= 0.03)^*^, V3 vs V4 (p= 0.002)^**^, V3 vs V5 (p= ns) and V4 vs V5 (p=0.03)^*^.
