## Supplementary material for "Persistently reduced humoral and cellular immune response following third SARS-CoV-2 mRNA vaccination in anti-CD20-treated multiple sclerosis patients": Figure 3 legend

Graph A: Linear regression analysis of antibody levels 2-4 weeks after third vaccination in seropositive patients with time interval from the last ocrelizumab infusion. r^2^= 0.03779, p= 0.4395.

Graph B: Linear regression of antibody levels 2-4 weeks after third vaccination in seropositive patients with time interval between second and third vaccine. r^2^=0.00097, p=0.9.

BAU/ml = Binding antibody unit per ml.
