## Supplementary material for "Persistently reduced humoral and cellular immune response following third SARS-CoV-2 mRNA vaccination in anti-CD20-treated multiple sclerosis patients": Figure 4 legend

Scatter plot graph depicting frequencies of spike-reactive CD4^+^ and CD8^+^ cells mean with SEM at visit 1 (V1), visit 3 (V3) and visit 5 (V5). Y-axis in two segments from 0-1 × 10^9^ cells/l and 2-6 × 10^9^ cells/l.

V1= Visit 1 (0-7 days before the first vaccine), V3 = Visit 3 (2-4 weeks after the second vaccination), and V5 = Visit 5 (2-4 weeks after the third vaccination).

AIM = Activation-induced markers of positive T-cells

SEM = standard error of the mean
